# Impact of COVID-19 epidemic curtailment strategies in selected Indian states: an analysis by reproduction number and doubling time with incidence modelling

**DOI:** 10.1101/2020.05.10.20094946

**Authors:** Arun Mitra, Abhijit P Pakhare, Adrija Roy, Ankur Joshi

**Author notes:** Corresponding author. Ankur Joshi, MD. Assistant Professor, Department of Community and Family Medicine, All India Institute of Medical Sciences, Bhopal, India.

## Abstract

The Government of India in network with the State Governments has implemented the epidemic curtailment strategies inclusive of case-isolation, quarantine and lockdown in response to ongoing novel coronavirus (COVID-19) outbreak. In this manuscript we attempt to estimate the effect of these steps across ten Indian states using crowd-sourced data. The chosen transmission parameters are -reproduction number (R0), doubling time and growth rate during the early epidemic phase (15 days into lockdown) and 30 days into lockdown (23rd April 2020) through maximum likelihood approach.

The overall analysis shows the decreasing trends in reproductive numbers and growth rate (with few exceptions) and incremental doubling time. The curtailment strategies employed by the Indian government seems to be effective in reducing the transmission parameters of the COVID-19 epidemic. The effective reproductive numbers are still higher above the threshold of 1, the resultant absolute numbers tend to augment as a function of time. The curtailment strategy thus may take into account these findings while formulating further course of actions.

## 1. Introduction

The World Health Organization (WHO) declared the Novel Coronavirus Outbreak (COVID-19) as a pandemic on 11th March 2020, calling for immediate action to be taken on by all countries in terms of stepping up treatment, detection, and reduction of transmission. As of 26th April 2020, a total of 2.96 million confirmed cases with over 200 thousand deaths have been reported in 185 countries.(1) The Ministry of Health and Family Welfare, Govt. of India reported over 20000 cases across 32 states/union territories with 872 deaths.(2) Government of India, initiated various Non-Pharmaceutical Intervention (NPI) which included social distancing measures like lockdown. Nationwide lockdown began from 24th March 2020 onwards. It had restrictions on non-essential travel, prohibition of mass gatherings etc.

In spite of assumed uniform susceptibility of Indian population to COVID-19, the trends till now are showing a variegated force of infection in different states. It is important to capture these state specific variations as they may offer important insights into the current mitigation strategies. This is specifically contextual in amidst of lockdown strategy adopted by the country to curtail the impact and flatten the peak of the COVID-19 epidemic.(3-5) the quantification of which may aid to plan future intervention strategies. The scope of this manuscript is to estimate the basic/effective reproduction number (R0) and doubling time at 15 days into the lockdown (early epidemic) and at day-30 of the lockdown to see the cumulative effect of curtailment strategies (inclusive of lockdown) in selected states. The 10 states reporting highest numbers of COVID-19 cases as on 23^rd^ April, 2020 were chosen for this analysis. The database used for the analysis is in open-domain at www.covid19india.org. R0 and doubling time are chosen in view of their primary role in reflecting the force, consistency and continuity of an infectious disease which is critically important in COVID-19.

## 2. Methods

### 2.1. Data Source

The data source is a crowd-sourced database maintained at www.covid19india.org/.(6) We used the data from the line-listing of the cases reported as positive for COVID-19. The data was iteratively and progressively accessed through the database in coherence with creation and improvement in analysis code. The last access to database was made on 1^st^ May, 2020. We truncated the data up to 23th April 2020 for the purpose of this study. This buffer period of 7 days offered some immunity against the possible delay to add the cases and our limitation to access the data in real time. Statistical software R, version 3.6.2 was used to perform all statistical analysis and model development.

### 2.2. Data preparation and analysis

The data is prepared for analysis in the following steps:

1. Loading the *.json file containing the raw line-list data
2. A data-frame is then created and variables of interest are selected
3. The imported cases are then coded in the following fashion:
  a. All cases with travel history outside the country before the lockdown are coded as imported cases
  b. All cases reported after 15 days of the lockdown (i.e. 9th April 2020) irrespective of their travel history are coded as local cases
4. Data from the top 10 states with highest number of cases was subset
5. Respective incidence objects were created based on the timeframes described below.

We divided the timeline of the epidemic into three phases, the first is the phase was before lockdown i.e. 25th March 2020). The second phase was early epidemic (15 days into the lockdown) and the third phase was until 30 days since lockdown. However, the transmission parameters before lockdown were not estimated due to certain constraints described in the supplementary materials (Suppl.Table.1). The second phase (15 days into the lockdown) was considered as the baseline for estimation of the transmission parameters.

We used the package incidence to model the incidence and estimate growth rate and doubling time and package R0 to estimate the reproduction number (R0) for different states. (7-9) The package projections was used to simulate the epidemic outbreaks and project their respective trajectories based on the state specific transmission parameters.(10)

### 2.3. Principle behind estimation of R0

The R0 was estimated through the maximum likelihood method based on chain-binomial models.(11,12) The serial interval for COVID-19 (gamma distribution assuming a mean ± SD of 4.4 ± 3.0 days) was used as time stamp for the estimation.(13) The reason for choosing this method is the flexibility to use the entire available data set instead of using observations at the beginning of epidemic only. This method uses the MCMC (Markov chain Monte Carlo) algorithms for sampling from the distribution. This method optimizes β and S0 from the sequence of binomial likelihood with the fundamental assumptions of conditional independence. The confidence intervals of the estimates were obtained by profiling.

#### Ethical considerations

As mentioned, the data is taken from crowdsourced database available in public domain. This database uses count data published in State Bulletins and Official Reports. This study didn’t involve any sort of primary data gathering in any forms. (Personal interview, questionnaire etc).

## 3. Results

A total of 23,040 COVID-19 cases have been reported in India as of 23rd April 2020 of which 20,590 cases (89.4%) were seen in the selected 10 states. The proportion of imported cases was less than 2% in all the 10 states.

Table 1 shows the demographics and key relevant statistics pertaining to COVID-19 epidemic of the chosen states (as of 23rd April 2020).

**Table 1:** State wise distribution of population, reported cases, cases and test per million

| State Name | Population (in Million)<sup>#</sup> | Reported cases | Reported mortalities | Recovered number | Case fatality rate | Recovery Rate | Cases reported per million | Tests performed per million | Positivity Rate |
| --- | --- | --- | --- | --- | --- | --- | --- | --- | --- |
| **Maharashtra** | 112.4 | 6427 | 282 | 840 | 4.39 | 13.07 | 57.18 | 794 | 7.21 |
| **Gujarat** | 60.4 | 2624 | 112 | 252 | 4.27 | 9.60 | 43.44 | 702 | 6.19 |
| **Delhi** | 16.8 | 2376 | 50 | 808 | 2.1 | 34.01 | 141.43 | 1819 | 7.77 |
| **Rajasthan** | 68.5 | 1964 | 28 | 451 | 1.43 | 22.96 | 28.67 | 1018 | 2.82 |
| **Madhya Pradesh** | 72.6 | 1687 | 93 | 203 | 5.51 | 12.03 | 23.24 | 338 | 6.87 |
| **Tamil Nadu** | 72.1 | 1683 | 20 | 752 | 1.19 | 44.68 | 23.34 | 915 | 2.55 |
| **Uttar Pradesh** | 199.8 | 1510 | 24 | 206 | 1.59 | 13.64 | 7.56 | 228 | 3.32 |
| **Telangana** | 35.2 | 970 | 25 | 252 | 2.58 | 25.98 | 27.56 | 425* | 6.48* |
| **Andhra Pradesh** | 49.5 | 893 | 27 | 141 | 3.02 | 15.79 | 18.04 | 970 | 1.86 |
| **West Bengal** | 91.3 | 456 | 15 | 79 | 3.29 | 17.32 | 4.99 | 88 | 5.71 |

<sup>#</sup> According to Census 2011 \*Testing data for 19th April 2020 was used

Figure 1, is a composite plot where lines diagram shows the trends of cumulative number of cases in reference to time and the histogram shows the proportional increase in cases/day for a specific state on that specific day. The two v-line divide the whole interface into before lockdown, early epidemic (15 days into lockdown) and current time frame (30 days into lockdown).

**Figure 1:**
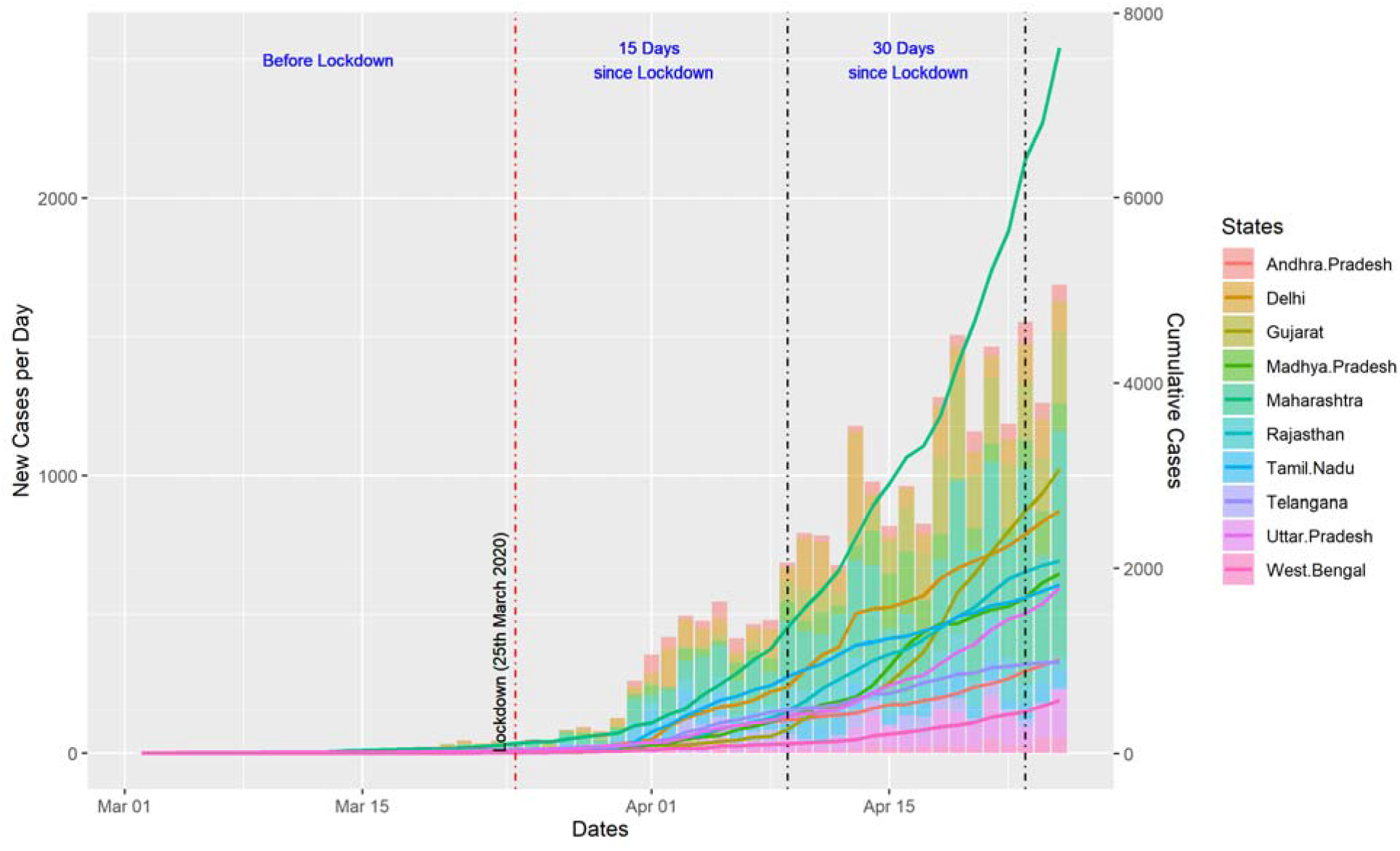
Date wise COVID-19 cases reported by states in India

### 3.1. Changes in reproductive number, doubling time and growth rate

Table 2 shows the baseline R0 (calculated at 15 days into lockdown) and effective R0 at 30 days into lockdown. The respective doubling time is also shown at these time points. The estimates in doubling time during the early epidemic in some states show high degree of unreliability with wide confidence intervals. This may be due to events following the Poisson process at the beginning of epidemic where the approximate average time between events is known yet the case to case timing varies significantly. All these estimates are shown as NA.

**Table 2:**
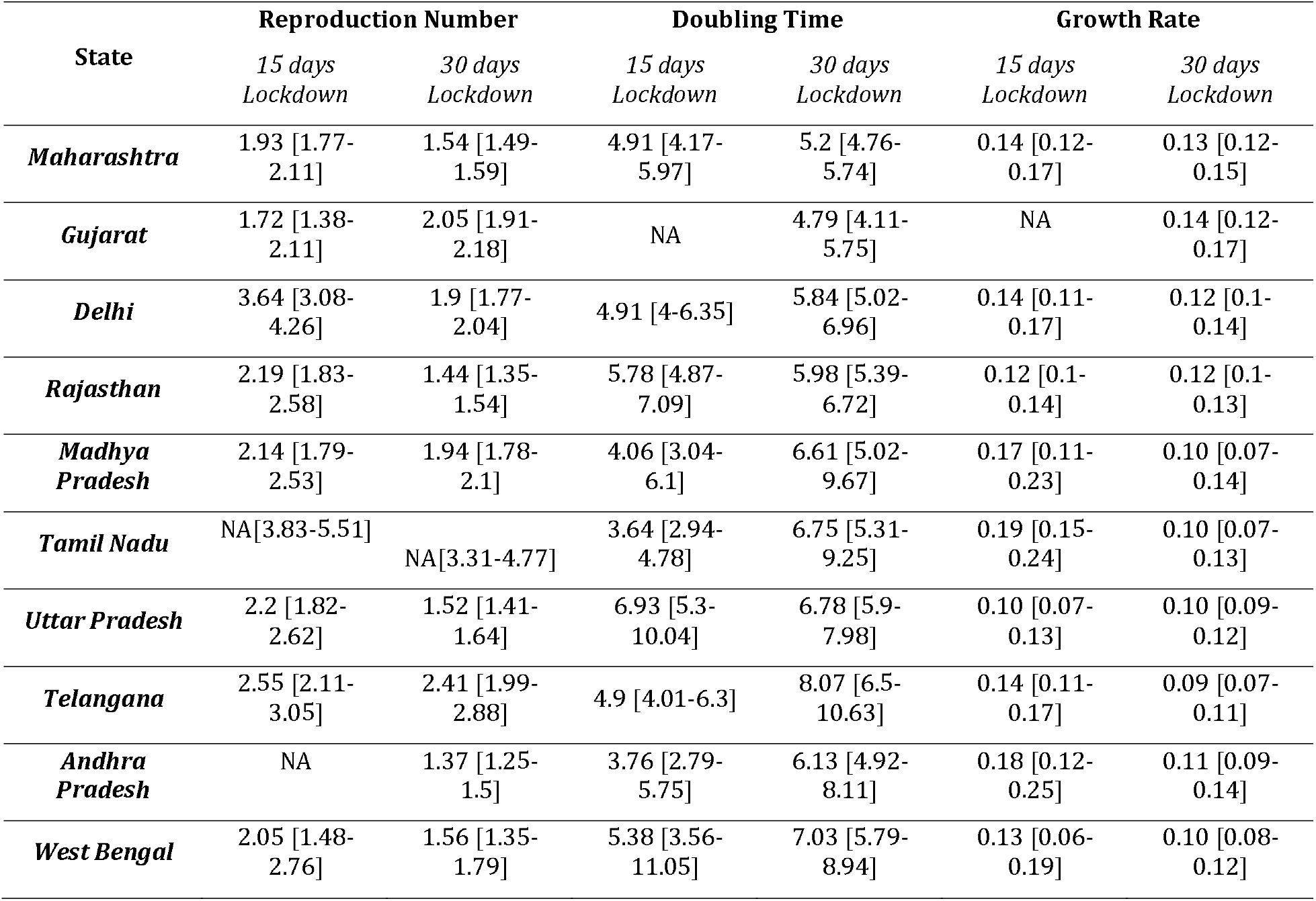
State-wise change in reproductive number, doubling time and growth rate over time

Seven of the ten selected states saw a reduction in reproduction number (R0) between the early epidemic phase and the current timeframe. Highest decrease in R0 was seen in Andhra Pradesh (73%) followed by Delhi (43%) and Rajasthan (30%).

Doubling time also changed with evolving outbreak. Increase in doubling time means slow growth rate of an outbreak. At least 5 states showed the substantial increase in doubling time. The growth rates of 8 of 10 states showed decline between the two-time intervals. Additional analysis is provided in the Supplementary Appendix. 1 along with the R Code.

### 3.2. Modeling Incidence & Future Projections

Regression of log-incidence over time was used to model the cumulative-incidence. The package ‘projections’ was used to simulate 1000 probable epidemic outbreak trajectories and plot the future daily cumulative incidence predictions based on probability mass function dependent branching process assuming it follows a Poisson distribution. This was done to curve-fit the robustness of R0 and check it by plotting against new incidence. The reproduction numbers of the third phase (i.e. 30 days into lockdown) were used to model the incidence and predict the cumulative case load for the selected states.

Amongst the 10-day projected cases, seven of the ten states had observed values within the predicted range. States of Rajasthan, Madhya Pradesh and Telangana observed lesser cases than predicted (Suppl.Table.2). Detailed description on the methods of projections is provided in the Supplementary Appendix. 2.

## 4. Discussion

This study evaluates impact of nation-wide lockdown on COVID-19 cases in ten states of India. At the beginning of the outbreak, states reported high transmissibility and lower doubling time. National lockdown was implemented from 24th March 2020. The effective R0 in several states has come down by the adopted curtailment strategies / lockdown compared to what was estimated at the beginning of the epidemic. As the final epidemics size’s relation with R0 is exponential and not linear, this reduction if sustained may considerably decrease the total number of affected persons compared to initial estimates. However, two factors should be considered at this moment. Firstly, effective reproduction numbers (R_t_) need to be further reduced in-order to flatten or change the direction of the epidemic curve it may be perceived state-wise variations in its magnitude. Secondly, although the doubling time has increased in relative terms, the epidemic still follows an exponential trajectory and the current daily incidence is much more as compared to beginning of the epidemic.

The overall picture suggests the initial success of Indian states to curtail the rise of curve. This reduction can mainly be explained by reduced number of contacts among people owing to movement restrictions. Studies on the impact of lockdown in other countries also reported reduction in reproductive number which translates in to flattening of the curve and delaying of peak.(14-19) Yet as mentioned earlier, the effective R0 estimations are dynamic and may change over age structure, time and nature of intervention. Continuing nationwide lockdown at this scale and with the same intensity may not be further possible for the government because of collateral implications in long term. Therefore, in the post-lockdown era, it might be a challenge to maintain this path and this may be the period where the absolute burden of the infected persons will be high.(20,21) There remains a higher reported probability of serious infections (including intensive care) in geriatric population compared to younger adults. (22) This coupled with the higher prevalence of comorbid conditions (~50%) in individuals over 60 years in India may warrant a strategy tailored to this section of population.(23) This also suggests that in addition to identification of infection, it is imperative to shift the focus on mortality prevention. Containment strategies like lockdown has given us the much-needed opportunity to delay the peak flatten the epidemic-curve. The time bought should be utilized to intensify the surveillance among ‘at-risk’ individuals and buttress the health infrastructure including hospital beds with oxygen availability and critical care beds with ventilators and telemedicine.(24-26)

The results of this study should be interpreted with certain caveats apart from the inherent limitations of crowd-sourcing nature of the data. The estimates might be influenced by certain effect modifiers and confounders like population density, climatic variations and violation of assumption of random mixing, yet the methods are robust in terms of conditional independence and MCMC methods used to tackle the Bayesian influence. Conceptually, this phenomenon is dynamic and non-linear in nature and hence should be read with caution

## Data Availability

Code of analysis is provided as supplementary file. running the code will fetch dataset from publicly available API.

## Acknowledgement

We would like to thank the team of COVID-19 India Tracker (www.covid19india.org) for their initiative in envisioning and developing the website. We are grateful to the members of the ‘Covid-19India.orgFolding@Home Team’ Telegram group for their invaluable and continued support without which this manuscript would not be possible.

## Supplementary Files

Supplementary Appendix-1: R-code for analysis

Supplementary Appendix-2: Incidence modelling and projections

Supplementary Table-1: Number of imported and local cases

Supplementary Table-2: Number of projected cases and actual cases

